# Prevalence and Clinical Profiling of Dysglycemia and HIV infection in Persons with Pulmonary Tuberculosis in Brazil

**DOI:** 10.1101/2021.10.29.21265663

**Authors:** María B. Arriaga, Mariana Araújo-Pereira, Beatriz Barreto-Duarte, Caio Sales, João Pedro Miguez-Pinto, Evelyn B. Nogueira, Betânia M. F. Nogueira, Michael S. Rocha, Alexandra B. Souza, Aline Benjamin, Jamile G. de Oliveira, Adriana S. R. Moreira, Artur T. L. Queiroz, Moreno M. S. Rodrigues, Renata Spener-Gomes, Marina C. Figueiredo, Betina Durovni, Solange Cavalcante, José R. Lapa-e-Silva, Afrânio L. Kristki, Marcelo Cordeiro-Santos, Timothy R. Sterling, Valeria C. Rolla, Bruno B. Andrade, the RePORT-Brazil consortium

## Abstract

**Background:** There are scarce data on the prevalence and disease presentation of HIV in patients with tuberculosis (TB) and dysglycemia (diabetes [DM] and prediabetes [PDM]), especially in TB-endemic countries.

**Methods:** We assessed the baseline epidemiological and clinical characteristics of patients with culture-confirmed pulmonary TB, enrolled in a multicenter prospective cohort in Brazil (RePORT-Brazil) during 2015-2019. Dysglycemia was defined by elevated glycated hemoglobin and stratified as PDM or DM. Additionally, we used data from TB cases obtained through the Brazilian National Notifiable Diseases Information System (SINAN), during 2015-2019. In SINAN, diagnosis of diabetes was based on self-report. Logistic regression models were performed to test independent associations between HIV, dysglycemia status, and other baseline characteristics in both cohorts.

**Results:** In the RePORT-Brazil cohort, the prevalence of DM and of PDM was 23.7% and 37.8%, respectively. Furthermore, the prevalence of HIV was 21.4% in the group of persons with TB-dysglycemia and 20.5% in that of patients with TBDM. In the SINAN cohort, the prevalence of DM was 9.2%, and among the TBDM group the prevalence of HIV was 4.1%. Logistic regressions demonstrated that aging was independently associated with PDM or DM in both the RePORT-Brazil and SINAN cohorts. In RePORT-Brazil, illicit drug use was associated with PDM, whereas a higher body mass index (BMI) was associated with DM occurrence. Of note, HIV was not associated with an increased risk of PDM or DM in patients with pulmonary TB in both cohorts. Moreover, in both cohorts, the TBDM-HIV group presented with a lower proportion of positive sputum smear and a higher frequency of tobacco and alcohol users.

**Conclusion:** There is a high prevalence of dysglycemia in patients with pulmonary TB in Brazil, regardless of the HIV status. This reinforces the idea that DM should be systematically screened in persons with TB. Presence of HIV does not substantially impact clinical presentation in persons with TBDM, although it is associated with more frequent use of recreational drugs and smear negative sputum samples during TB screening.

## 1 Introduction

Approximately one-quarter of the world population is thought to be infected with *Mycobacterium tuberculosis* (Mtb) and about 5%-10% of those will develop active disease at some point in their lives, which represents a substantial public health problem (World Health Organization, 2021). Several factors are related to the development of active tuberculosis (TB), such as immunological, genetic, and metabolic factors. Importantly, metabolic disorders associated with glycemic status are considered important risk factors for the development active TB and also for unfavorable anti-TB treatment outcomes (Hensel et al., 2016). In addition, the immune deterioration caused by human immunodeficiency virus (HIV) favors the multiplication of Mtb and the progression to active TB (He et al., 2020).

Dysglycemia is a spectrum of metabolic dysfunctions related to glucose metabolism in the body, which includes several diseases, especially prediabetes (PDM) and diabetes (DM) (American Diabetes, 2021). Approximately 422 million people worldwide live with DM, most of whom are in low-and middle-income countries. Likewise, a significant part of the world’s population suffers from PDM, an intermediate state of insulin resistance that partially affects the entry of glucose into cells (Abdul-Ghani et al., 2006;Goncalves et al., 2019). Interestingly, TB is similarly focused on low-and middle-income countries, which is a problem as DM triples the risk of developing active TB (Hayashi and Chandramohan, 2018). Furthermore, 15.3% of people with TB worldwide have DM as a comorbidity (Noubiap et al., 2019). Persons with TB-DM usually exhibit a different clinical presentation, which includes higher frequency of extensive or cavitary pulmonary TB, a higher bacillary load in sputum and delayed mycobacterial clearance compared to normoglycemic TB patients (Singla et al., 2006;Gil-Santana et al., 2016). Although much has been described on the interaction between TB and DM in different settings, most of the studies investigated a limited number of participants, and larger studies are warranted to better define such interactions. In addition, the clinical outcomes as well as the pathophysiological mechanisms of patients with TB-DM are still poorly understood (Jeon and Murray, 2008;Hayashi and Chandramohan, 2018).

In addition to the importance of metabolic disorders, conditions that directly affect the immune response against TB are also a relevant problem as they contribute to more severe manifestations (World Health Organization, 2020). Importantly, people living with HIV (PLWH) are approximately 50 times more likely to develop active TB than those without HIV exposure (World Health Organization, 2020). Moreover, in 2019, PLWH accounted for 1.2 million (8.2%) of the approximately 10 million people with TB worldwide and of those, 208,000 deaths were related to HIV comorbidity (World Health Organization, 2020). On the other hand, persons living with both TB and HIV often experience accelerated HIV disease progression and TB is placed as the most common opportunistic infection inducing high morbidity (World Health Organization, 2020). HIV has been shown to modify the course of TB by causing severe immunosuppression and Mtb dissemination to multiple organs and increased mortality (Geldmacher et al., 2010;Moir et al., 2011).

Brazil has a high burden of TB-DM (Noubiap et al., 2019) and TB-HIV (World Health Organization, 2021). Despite of the high of these comorbidities, to our knowledge there is no information that explores in detail the association between HIV and TB-DM and its impact on clinical presentation of affected persons in the country. The scarce information that exists come from studies performed in African populations and with results that are not consistent with each other (Bailey and Ayles, 2017;Oni et al., 2017). Because of the abovementioned reasons, studies that examine the overlap of metabolic and immunological diseases associated with TB are needed to better understand the spectrum of disease presentation of patients with multiple comorbidities such as TB-DM-HIV. In the present study, we aimed at contributing to fill this gap in knowledge in the context of TB, dysglycemia, and HIV-infection, through the identification and characterization of HIV prevalence and its association with glycemic status among persons with active pulmonary TB, in the Regional Prospective Observational Research in Tuberculosis (RePORT-Brazil) study, which is a large multicenter prospective cohort of culture-confirmed pulmonary TB persons which has been shown to be representative of the TB cases reported in the Brazilian national TB registry (Hamilton et al., 2015;Arriaga et al., 2021). We also investigated such associations in TB cases reported to the Brazilian National TB Registry through the National System of Diseases Notification (SINAN).

## 2 METHODS

### 2.1 Ethics Statement

All clinical investigations were conducted according to the principles of the Declaration of Helsinki. The RePORT-Brazil protocol, informed consent, and study documents were approved by the institutional review boards at each study site and at Vanderbilt University Medical Center (CAAE: 25102414.3.2009.5543). Participation in RePORT-Brazil was voluntary, and written informed consent was obtained from all such participants.

### 2.2 Study design – RePORT-Brazil

This was a multicenter prospective observational cohort study of individuals ≥ 18 years old with culture-confirmed pulmonary TB. RePORT-Brazil study sites are located in Manaus (Amazonas state, Northern region), Salvador (Bahia state, Northeastern region), and Rio de Janeiro (Rio de Janeiro state, Southeastern region), with a total of five health units: Instituto Nacional de Infectologia Evandro Chagas, Clínica da Família Rinaldo Delamare, and Secretaria Municipal de Saúde de Duque de Caxias (Rio de Janeiro), Instituto Brasileiro para Investigação da Tuberculose, Fundação José Silveira (Bahia), and Fundação de Medicina Tropical Doutor Heitor Vieira Dourado (Amazonas), representing both a heterogeneous population and the Brazilian cities with highest TB burden (Hamilton et al., 2015;Arriaga et al., 2021).

### 2.3 Data collection – RePORT-Brazil

Between 2015 and 2019, TB cases were interviewed for sociodemographic, clinical and epidemiological data such as age, sex, race/ethnicity (self-reported), body mass index (BMI), income, smoking status, passive smoking status (living with someone who smokes), alcohol and illicit drug use, and clinical data such as presence of TB symptoms (cough, fever, weight loss, fatigue, night sweats, chest pain) and had the following tests performed: chest X-ray, HIV serologic test (the test was not performed if the individuals had a previous diagnosis of HIV), CD4 and viral load (if HIV serology was positive or previous diagnosis of HIV-infection), complete blood count, glycated hemoglobin (HbA1c), sputum smear microscopy, Xpert-MTB-RIF (if available) and mycobacterial culture (Lowenstein-Jensen medium or BD BACTEC MGIT). Patients who received TB treatment or fluoroquinolones for >7 days in the 30 days prior to TB diagnosis and pregnant women were excluded. We only analyzed information collected at the study baseline.

### 2.4 Notifiable diseases information system (SINAN), Brazilian Ministry of Health

SINAN is a system for the notification of transmissible diseases, including TB, that has been implemented, supported, and maintained by the Brazilian Ministry of Health (Ministério da Saúde do Brasil and Secretaria de Vigilância em Saúde, 2020). Data were collected from TB patients ≥ 18 years old with information about “diabetes status”, between 2015 and 2019. Persons who were homeless, prisoners, pregnant, or had extrapulmonary TB were excluded, resulting in a population of 279,143 individuals. TB was diagnosed according to the Brazilian Ministry of Health criteria, detailed in the Manual of Recommendations for the TB Control in Brazil (Ministério da Saúde do Brasil and Secretaria de Vigilância em Saúde, 2013). After TB diagnosis, the information collected at the baseline and the laboratory results were recorded on a standardized form that, individual characteristics (sex, age, race, education level, alcohol consumption, illicit drug use, smoking habits, and comorbidities), the presence of DM condition (“yes” or “no” options) and HIV-infection, among others (Ministério da Saúde do Brasil and Secretaria de Vigilância em Saúde, 2013).

### 2.5 Study definitions

In pulmonary TB cases from RePORT-Brazil, participants with HbA1c ≥ 5.7% were classified as dysglycemic and those with HbA1c < 5.7% were considered normoglycemic. Study participants were also classified as having DM (HbA1c ≥ 6.5%), PDM (HbA1c= 5.7-6.4%) or normoglycemia (HbA1c< 5.7%), following American Diabetes Association (ADA) guidelines (International Diabetes Federation, 2019).

### 2.6 Data analysis

Categorical variables were presented as proportions and compared using a two-sided Pearson’s chi-square test (with Yates’s correction) or Fisher’s two-tailed test in 2×3 or 2×2 tables, respectively. Continuous variables were presented as median and interquartile range (IQR) and compared using the Mann Whitney *U* (between 2 groups) or Kruskal Wallis test (between ≥2 groups). Viral load values and CD4 count were transformed to log10 for analyses. Multinomial and binomial logistic regression models with stepwise method (Wald) were performed to evaluate independent associations between clinical characteristics of pulmonary TB cases and presence of diabetes and/or prediabetes in the Report-Brazil and SINAN cohorts. Parameters with p-values ≤ 0.2 in univariate analyses were included in multivariable models. P-values < 0.05 were considered statistically significant. All the analyses were pre-specified. Statistical analyses were performed using SPSS 24.0 (IBM statistics), Graphpad Prism 9.0 (GraphPad Software, San Diego, CA) and R 3.1.0 (R Foundation, Austria).

## 3 RESULTS

### 3.1 Characteristics of the study participants

RePORT-Brazil enrolled 1,162 patients with culture-positive pulmonary TB during 2015-2019 from the five centers of the consortium. The prevalence of dysglycemia at TB diagnosis was 61.5% (95%CI: 58.6 – 64.2). Compared to normoglycemic individuals, those with dysglycemia were more likely male (68.8% vs 61.8%, p =0.018) and older (39, IQR:29-52 years, p< 0.001). Among TB-DM cases, 122/275 (44.4%) had previous diagnosis of DM. The dysglycemia group also exhibited higher frequency of self-reported *pardo* race (n=388, 54.4%, p=0.007), a higher median of BMI value (20.5, IQR:18.4-23.1; p<0.001) and a higher frequency of self-reported weight loss (n=597, 93%; p=0.016) but not of other TB symptoms (**Table 1**).

**Table 1.**
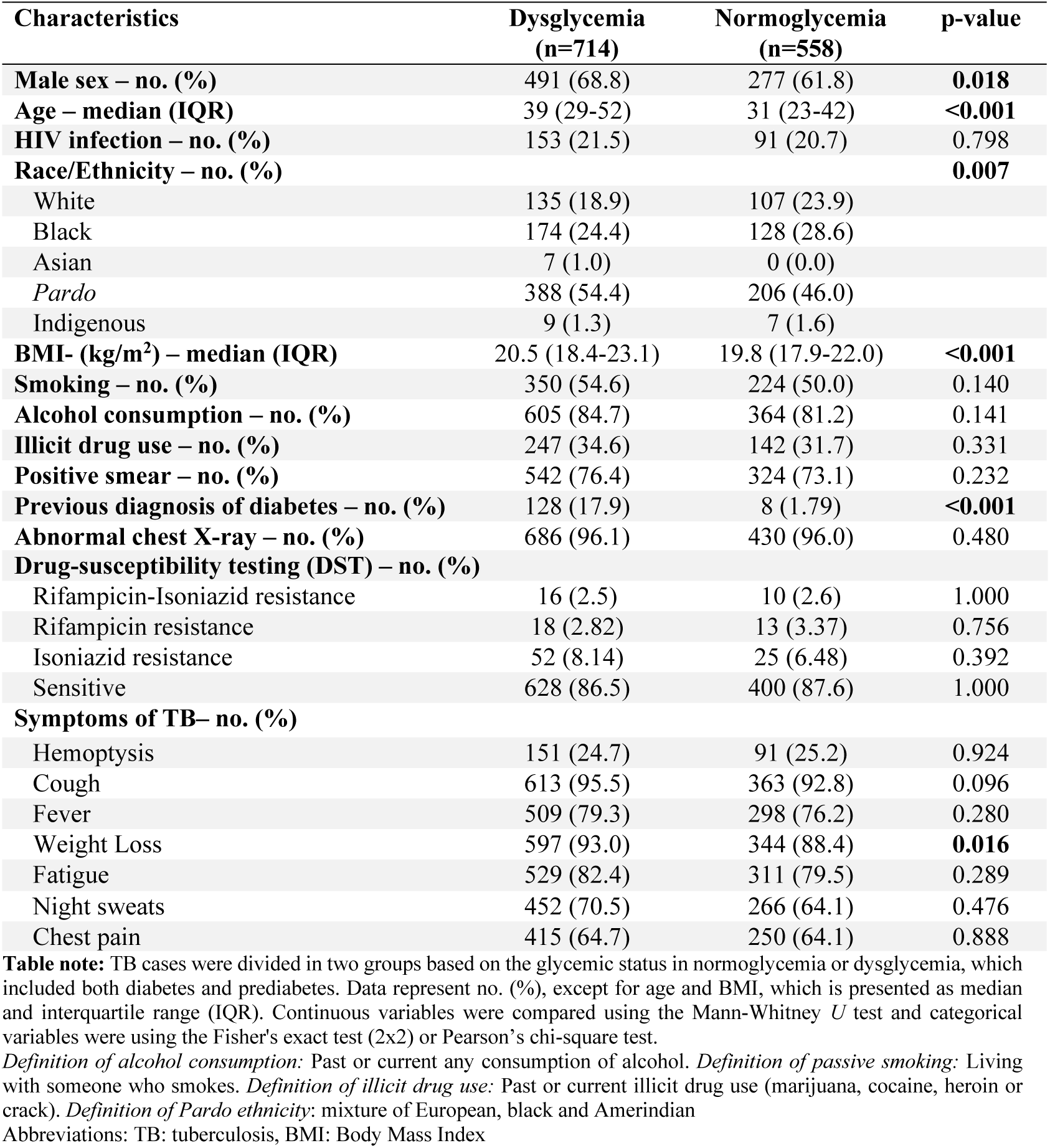
Characteristics of TB cases by glycemic status in RePORT-Brazil cohort.

| Characteristics | Dysglycemia<br>(n=714) | Normoglycemia<br>(n=558) | p-value |
| --- | --- | --- | --- |
| Male sex – no. (%) | 491 (68.8) | 277 (61.8) | **0.018** |
| Age – median (IQR) | 39 (29-52) | 31 (23-42) | **<0.001** |
| HIV infection – no. (%) | 153 (21.5) | 91 (20.7) | 0.798 |
| Race/Ethnicity – no. (%) |  |  | **0.007** |
| White | 135 (18.9) | 107 (23.9) |  |
| Black | 174 (24.4) | 128 (28.6) |  |
| Asian | 7 (1.0) | 0 (0.0) |  |
| Pardo | 388 (54.4) | 206 (46.0) |  |
| Indigenous | 9 (1.3) | 7 (1.6) |  |
| BMI- (kg/m<sup>2</sup>) – median (IQR) | 20.5 (18.4-23.1) | 19.8 (17.9-22.0) | **<0.001** |
| Smoking – no. (%) | 350 (54.6) | 224 (50.0) | 0.140 |
| Alcohol consumption – no. (%) | 605 (84.7) | 364 (81.2) | 0.141 |
| Illicit drug use – no. (%) | 247 (34.6) | 142 (31.7) | 0.331 |
| Positive smear – no. (%) | 542 (76.4) | 324 (73.1) | 0.232 |
| Previous diagnosis of diabetes – no. (%) | 128 (17.9) | 8 (1.79) | **<0.001** |
| Abnormal chest X-ray – no. (%) | 686 (96.1) | 430 (96.0) | 0.480 |
| Drug-susceptibility testing (DST) – no. (%) |  |  |  |
| Rifampicin-Isoniazid resistance | 16 (2.5) | 10 (2.6) | 1.000 |
| Rifampicin resistance | 18 (2.82) | 13 (3.37) | 0.756 |
| Isoniazid resistance | 52 (8.14) | 25 (6.48) | 0.392 |
| Sensitive | 628 (86.5) | 400 (87.6) | 1.000 |
| Symptoms of TB– no. (%) |  |  |  |
| Hemoptysis | 151 (24.7) | 91 (25.2) | 0.924 |
| Cough | 613 (95.5) | 363 (92.8) | 0.096 |
| Fever | 509 (79.3) | 298 (76.2) | 0.280 |
| Weight Loss | 597 (93.0) | 344 (88.4) | **0.016** |
| Fatigue | 529 (82.4) | 311 (79.5) | 0.289 |
| Night sweats | 452 (70.5) | 266 (64.1) | 0.476 |
| Chest pain | 415 (64.7) | 250 (64.1) | 0.888 |

**Table note:** TB cases were divided in two groups based on the glycemic status in normoglycemia or dysglycemia, which included both diabetes and prediabetes. Data represent no. (%), except for age and BMI, which is presented as median and interquartile range (IQR). Continuous variables were compared using the Mann-Whitney *U* test and categorical variables were using the Fisher's exact test (2x2) or Pearson's chi-square test.
*Definition of alcohol consumption:* Past or current any consumption of alcohol. *Definition of passive smoking:* Living with someone who smokes. *Definition of illicit drug use:* Past or current illicit drug use (marijuana, cocaine, heroin or crack). *Definition of Pardo ethnicity:* mixture of European, black and Amerindian
Abbreviations: TB: tuberculosis, BMI: Body Mass Index

### 3.2 Characteristics of TB cases by glycemic status

In RePORT-Brazil, the DM and PDM prevalence at TB diagnosis was 23.7% (95%CI: 21.31-26.2%) (n=275), and 37.8% (95%CI: 35.0%-40.6%) (n=439), respectively (**Figure 1A**). Several clinical characteristics differed between normoglycemic and dysglycemic TB patients, with significant differences in frequency of sex (p=0.027), age (p<0.001) race/ethnicity (p<0.001), BMI values (p<0.001) and the frequency of self-reported weight loss as a symptom (p=0.033) between the three groups (**Table 2**).

**Table 2.** Characteristics of TB cases by DM status in RePORT-Brazil cohort.

| Characteristics | Diabetes<br>(n=275) | Prediabetes<br>(n=439) | Normoglycemia<br>(n=448) | p-value |
| --- | --- | --- | --- | --- |
| **Sex male – no. (%)** | 182 (66.2) | 309 (70.4) | 277 (61.8) | **0.027** |
| **Age – median (IQR)** | 46 (36-55) | 36 (26-47) | 31 (23-42) | **<0.001** |
| **HIV infection – no. (%)** | 56 (20.5) | 97 (22.2) | 91 (20.7) | 0.821 |
| **Race/Ethnicity – no. (%)** |  |  |  | **<0.001** |
| White | 43 (15.7) | 92 (21.0) | 107 (23.9) |  |
| Black | 58 (21.2) | 116 (26.4) | 128 (28.6) |  |
| Asian | 2 (0.73) | 5 (1.14) | 0 (0.0) |  |
| *Pardo* | 167 (60.9) | 221 (50.3) | 206 (46.0) |  |
| Indigenous | 4 (1.46) | 5 (1.14) | 7 (1.56) |  |
| **BMI (kg/m<sup>2</sup>)-median (IQR)** | 21.6 (19.1-24.4) | 19.9 (18.4-21.8) | 19.8 (17.9-22.0) | **<0.001** |
| **Smoking – no. (%)** | 158 (57.6) | 232 (52.8) | 224 (50.0) | 0.150 |
| **Alcohol consumption – no. (%)** | 242 (88.0) | 363 (82.7) | 364 (81.2) | 0.053 |
| **Illicit drug use – no. (%)** | 77 (28.0) | 170 (38.8) | 142 (31.7) | **0.007** |
| **Positive Smear – no. (%)** | 220 (80.3) | 332 (74.0) | 324 (73.1) | 0.077 |
| **Previous diagnosis of diabetes – no. (%)** | 122 (44.4) | 6 (1.4) | 8 (1.8) | **<0.001** |
| **Abnormal chest X-ray– no. (%)** | 269 (97.8) | 417 (95.0) | 430 (96.0) | 0.168 |
| **Drug-susceptibility testing (DST) – no. (%)** |  |  |  |  |
| Rifampicin resistance | 10 (4.0) | 8 (2.1) | 13 (3.4) | 0.331 |
| Isoniazid resistance | 21 (8.4) | 31 (8.0) | 25 (6.5) | 0.608 |
| Rifampicin-Isoniazid resistance | 8 (3.2) | 8 (2.1) | 10 (2.6) | 0.666 |
| Sensitive | 236 (84.4) | 392 (87.8) | 400 (87.5) | 1.000 |
| **Symptoms of TB– no. (%)** |  |  |  |  |
| Hemoptysis | 64 (27.2) | 87 (23.1) | 91 (25.2) | 0.515 |
| Cough | 235 (94.0) | 378 (96.4) | 363 (92.8) | 0.083 |
| Fever | 196 (78.4) | 313 (79.8) | 298 (76.2) | 0.466 |
| Weight Loss | 235 (94.0) | 362 (92.3) | 344 (88.4) | **0.033** |
| Fatigue | 214 (85.6) | 315 (80.4) | 311 (79.5) | 0.131 |
| Night sweats | 174 (69.9) | 278 (70.9) | 266 (68.2) | 0.708 |
| Chest paint | 165 (66.3) | 250 (63.8) | 250 (64.1) | 0.796 |

**Table note:** Data represent no. (%), except for age and BMI, which is presented as median and interquartile range (IQR). Continuous variables were compared using the Kruskal-Wallis test and categorical variables were using the Pearson's chi-square test. Bold values represent statistically significant.
*Definition of alcohol consumption:* Past or current any consumption of alcohol. *Definition of smoking:* Past or current cigarette smoker. *Definition of passive smoking:* Living with someone who smokes. *Definition of illicit drug use:* Past or current illicit drug use (marijuana, cocaine, heroin or crack)
*Definition of persistence of symptoms:* Patients who in the initial evaluation interview (baseline) reported indicated symptom and in the evaluation of visit 2 (month 2) still reported having such symptom.
*Definition of Pardo ethnicity:* mixture of European, black and Amerindian

**Figure 1.**
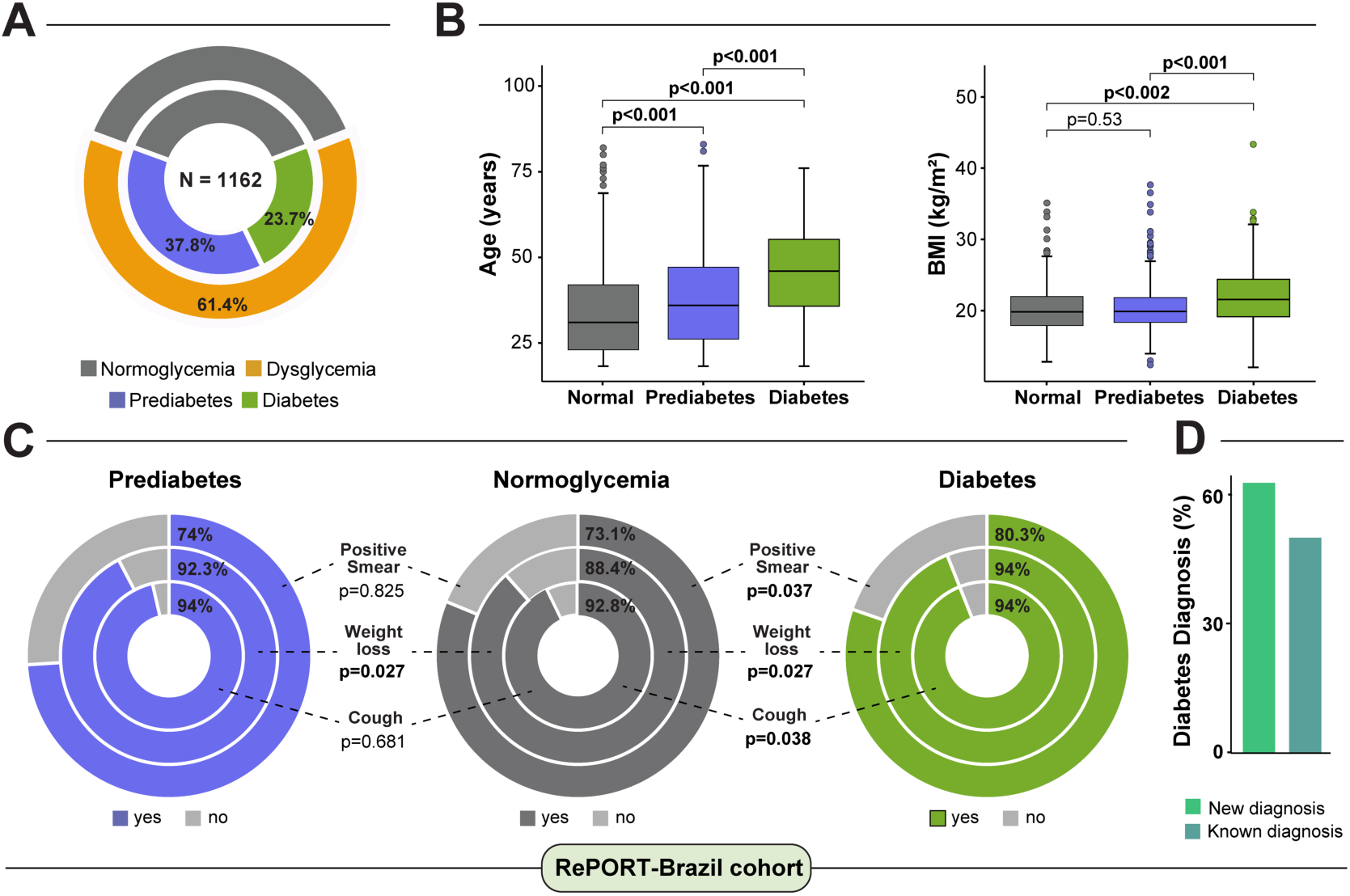
Characteristics of patients with pulmonary TB according to dysglycemia status in RePORT-Brazil cohort. **(A)** Among all the individuals with active pulmonary TB (n=1162), 61.4% had dysglycemia: 37.8% pre-DM and 23.7% DM. **(B)** Comparison of Age and BMI between groups were performed using Mann-Whitney *U* test. **(C)** Characteristics of the pulmonary TB cases stratified according to the presence of diabetes or prediabetes were compared with those from patients with normoglycemia using the Fisher’s exact test (additional comparisons are displayed in **Table 2**). **(D)** Frequency of new cases of DM diagnosis. The statistical analyzes were carried out only with the available data, omitting the cases with missing information. Abbreviations: TB: tuberculosis, BMI: Body Mass Index.

To evaluate these differences in more detail, we performed pair-wise comparisons between the groups. The highest median age was observed in the DM group (46 years; IQR: 36-55), which was significantly higher than PDM (36, IQR: 26-47) and normoglycemia (31, IQR: 23-42). We also observed differences between PDM and normoglycemia, with p<0.001 in both comparisons (**Figure 1B**). In addition, the DM group had higher median BMI (21.6, IQR: 19.1 - 24.4) than PDM (19.9, IQR: 18.4 - 21.80, p<0.001) and normoglycemia (19.8, IQR: 17.9 - 22.0, p<0.001), but there was no difference between PDM and normoglycemic individuals (p=0.53) (**Figure 1B**). TB patients with DM more frequently presented with positive smear (p=0.037), weight loss (p=0.027) and cough (p=0.038) than those with normoglycemia (**Figure 1C**). Patients with PDM similarly exhibited higher frequency of weight loss (p=0.027) compared with persons with normoglycemia at baseline (**Figure 1C**). Of note, 44.4% (n=122) of the participants with DM already knew about their diagnosis of DM before being enrolled in the study (**Figure 1D, Table 2**).

To assess whether the results obtained from the analyses of the RePORT-Brazil cohort mirrored the data from the overall Brazilian TB population, we characterized the TBDM cases reported to the SINAN registry (**Figure 2**). Of 279,143 pulmonary TB cases reported between 2015 and 2019, 25,765 had DM (self-reported), resulting in a prevalence of 9.2% (95%CI: 9.1%-9.3%) (**Table 3, Figure 2A**). Patients with TB-DM were older (55 years, IQR:46-64) than normoglycemic patients (40 years, IQR:29-54; p<0.001) (**Figure 2B**), had a higher frequency of abnormal chest X-ray (p<0.001), positive smear (p<0.001), positive culture (p<0.001) and were new cases more frequently reported (p <0.001) than in normoglycemic patients (**Figure 2C, Table 3**). In contrast, normoglycemic persons were more frequently men (p<0.001), reported greater consumption of alcohol (p<0.001) and illegal drug use (p<0.001) and more frequent tobacco use (p<0.001) than those with DM. Finally, normoglycemic TB patients were mainly black/*pardo* (p<0.001) and more frequently had drug-sensitive TB than TB-DM participants (p<0.001) (**Table 3**).

**Table 3.** Characteristics of TB cases by DM status in SINAN cohort.

| Characteristics | Diabetes<br>(n=25765) | Normoglycemia<br>(n=253378) | p-value |
| --- | --- | --- | --- |
| **Male sex – no. (%)** | 16172 (62.8) | 171638 (67.7) | **<0.001** |
| **Age – median (IQR)** | 55.0 (46.0-64.0) | 40.0 (29.0-54.0) | **<0.001** |
| **HIV infection – no. (%)** | 1051 (4.08) | 31073 (12.3) | **<0.001** |
| **ART use – no. (%)** | 484 (11.2) | 14175 (27.8) | **<0.001** |
| **Race/Ethnicity – no. (%)** |  |  | **<0.001** |
| White | 8243 (33.8) | 77442 (32.4) |  |
| Black/*Pardo* | 15750 (64.6) | 156749 (65.5) |  |
| Indigenous | 181 (0.74) | 2997 (1.25) |  |
| Asian | 210 (0.86) | 1953 (0.82) |  |
| **Abnormal chest X-ray – no. (%)** | 20113 (79.3) | 190895 (76.7) | **<0.001** |
| **Alcohol consumption – no. (%)** | 3820 (14.9) | 45548 (18.0) | **<0.001** |
| **Illicit drug use – no. (%)** | 1116 (4.35) | 26696 (10.6) | **<0.001** |
| **Smoking – no. (%)** | 4526 (17.7) | 47877 (19.0) | **<0.001** |
| **Positive smear – no. (%)** | 14649 (56.9) | 128692 (50.8) | **<0.001** |
| **Positive culture – no. (%)** | 4527 (17.6) | 42879 (16.9) | **0.003** |
| **Drug-susceptibility testing (DST) –**<br>**%** |  |  | 0.116 |
| Rifampicin resistance | 40 (1.8) | 428 (2.1) |  |
| Isoniazid resistance | 117 (5.4) | 1048 (5.1) |  |
| Rifampicin-Isoniazid resistance | 113 (5.2) | 857 (4.2) |  |
| Sensitive | 1906 (87.6) | 18211 (88.6) |  |

**Table note:** Data represent no. (%), except for age, which is presented as median and interquartile range (IQR). Continuous variables were compared using the Mann-Whitney *U* test and categorical variables were using the Fisher's exact test (2x2) or Pearson's chi-square test. Bold values represent statistically significant
*Definition of Pardo ethnicity:* mixture of European, black and Amerindian
Abbreviations: TB: tuberculosis, ART: antiretroviral therapy

**Figure 2.**
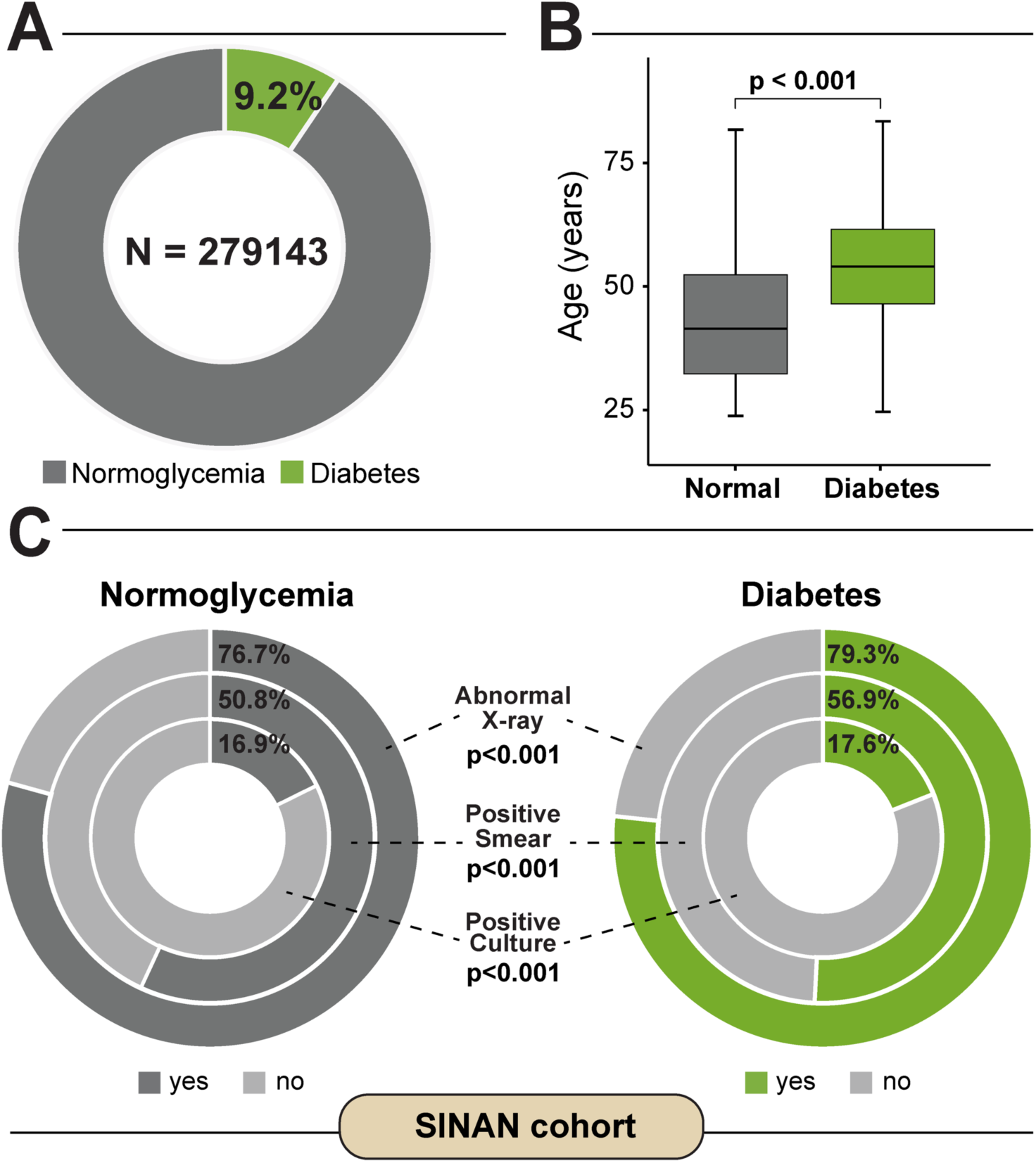
Characteristics of patients with pulmonary TB according to DM status in SINAN-TB cohort. **(A)** Among all the individuals with active pulmonary TB (n=287581) between 2015 and 2019 in Brazil, 9.2% had DM. **(B)** Comparison age between groups were performed using Mann-Whitney *U* test. **(C)** Characteristics of the pulmonary TB cases stratified according to the presence of diabetes were compared with those from patients with normoglycemia using the Fisher’s exact test (additional comparisons are displayed in **Table 2**). (Additional comparisons are displayed in **Table 3**). The statistical analyzes were carried out only with the available data, omitting the cases with missing information.

### 3.3 Characteristics of HIV status among TB cases with dysglycemia

In RePORT-Brazil, the association between HIV-infection status and dysglycemia at baseline in participants with active pulmonary TB was analyzed according to age, presence of DM or PDM as well as to HbA1c levels (**Figure 3**). Importantly, the distribution of HbA1c values among persons with TB did not differ significantly according to HIV-infection status (**Figure 3A**, left panel). In fact, HIV-infection was present in the minority of the active TB cases in all sub-categories of glycemic status. The HIV-infection prevalence in the TB-dysglycemia group was 21.4% (95%CI: 18.6%-4.3%) (**Figure 3A**, right panel). There was a significant difference in the frequency of TB patients with either DM (p<0.001) or HIV-infection (p<0.001) according to age category (**Figure 3B**), whereas there was no significant difference in the distribution of PDM among the different age categories (p=0.099). (**Figure 3B**). Of note, the subgroup of older participants (>48 years-old) exhibited the highest frequency of DM (**Figure 3B**).

**Figure 3.**
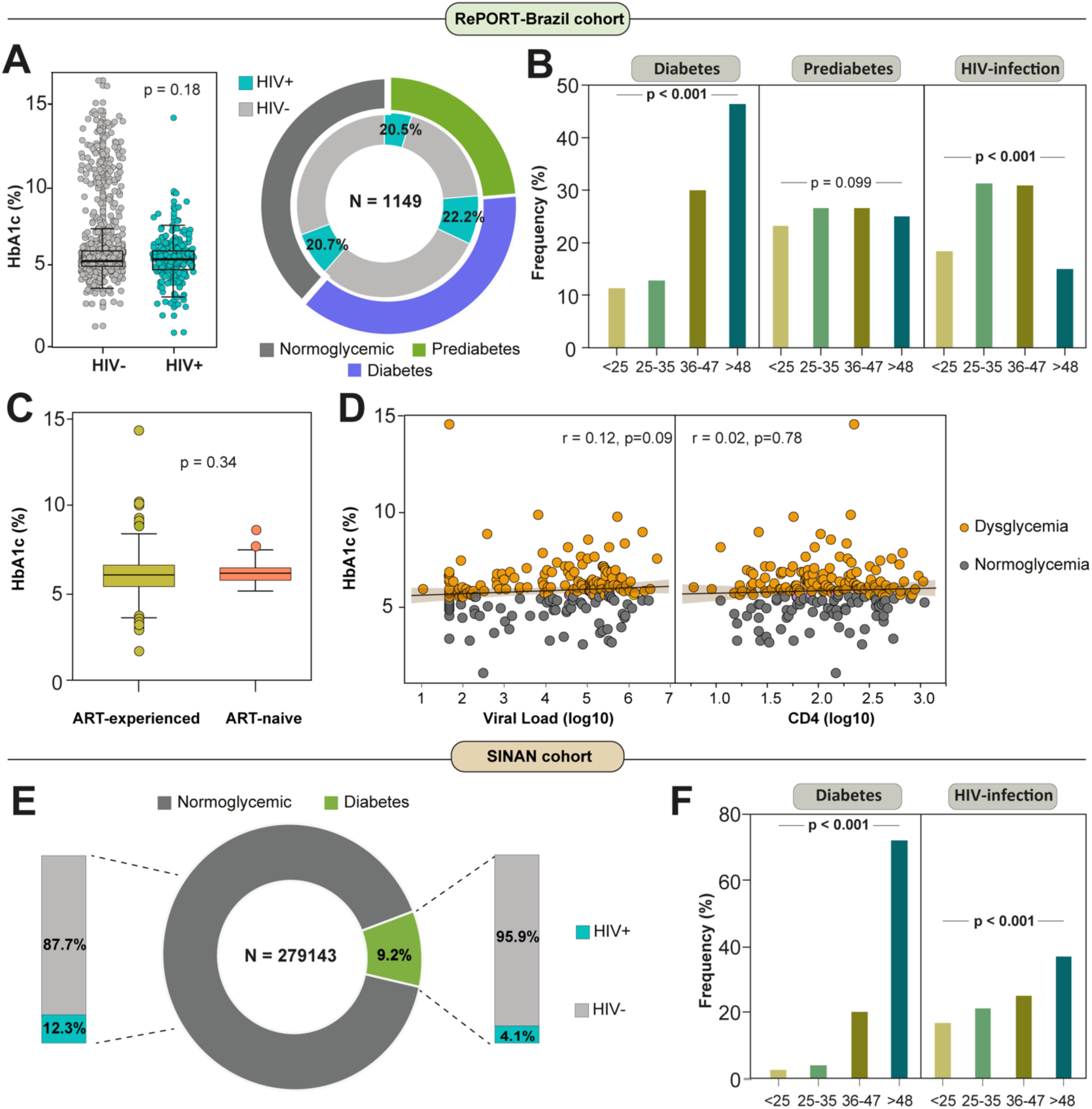
HIV infection among dysglycemic patients with active tuberculosis in RePORT-Brazil and SINAN cohorts. **(A)** Left panel: Scatter plot comparing distribution of HbA1c levels between subgroups of pulmonary TB cases per HIV infection status. Data were compared using the Mann-Whitney *U* test. Left panel: Total frequency of HIV infection among diabetic TB patients was 20.5%, among prediabetics was 22·2% and among normoglycemic patients was 20.7% (chi-square test p>0·05). **(B)** Frequency of individuals with diagnosis of diabetes, prediabetes and HIV infection in the indicated age category (in years) among pulmonary TB patients is shown. Data were compared using the Pearson’s chi-square test. **(C)** Box plot comparing distribution of HbA1c levels between subgroups of pulmonary TB cases per ART-experience and ART-naive status. Data were compared using the Mann-Whitney *U* test. **(D)** Spearman correlation between HbA1c and viral load (log10) levels (Left panel) and CD4 (log10) levels (Right panel) at baseline in pulmonary TB patients grouped according to the dysglycemic status. Line and shaded area represent linear curve fit with 95% confidence interval. **(E)** Total frequency of HIV infection among diabetic TB patients was 4.1% and among normoglycemic patients was 12.3% (chi-square test p>0·05). The statistical analyzes were carried out only with the available data, omitting the cases with missing information (14 patients were removed due to lack in HIV status). **(F)** Frequency of individuals with diagnosis of diabetes and HIV infection in the indicated age category (in years) among pulmonary TB patients is shown. Data were compared using the Pearson’s chi-square test. Abbreviations: ART: antiretroviral therapy.

Further comparisons revealed no differences in the distribution of HbA1c values between PLWH undertaking antiretroviral therapy (ART) and those who were ART-naïve at the time of study enrollment (**Figure 3C**). There was a non-significant positive correlation between HbA1c levels and HIV viral load (**Figure 3D**, left panel) and also between HbA1c concentrations and CD4 T-cell counts (**Figure 3D**, right panel) when all PLWH were considered regardless of the glycemic status (**Figure 3D**, left panel). In contrast, in the SINAN cohort, we found a prevalence of HIV-infection in the DM sub-group of 4.1% (95% CI: 3.8%-4.3%) (**Figure E**), lower than what was observed in the RePORT-Brazil cohort. Moreover, older TB patients (age >48 years) were more frequently found in in the DM and HIV-infection subgroups (p<0.001) than other age groups (**Figure F**).

### 3.4 Factors associated with dysglycemia in patients with active pulmonary TB

Multinomial logistic regression analyses were performed to test associations between characteristics of active pulmonary TB patients and the presence of PDM or DM in RePORT-Brazil participants. Results demonstrated that increases in age (per 1-year increase) were independently associated with an increased odds of PDM (adjusted odds ratio [aOR]: 1.02, IQR:1.01-1.03, p<0.001) or DM (aOR:1.06, IQR:1.04-1.07, p<0.001). Furthermore, self-reported illicit drug use (aOR:1.74, IQR:1.23-2.45, p=0.002) was related to increased odds of PDM but not DM. Higher BMI values (per 1Kg/m^2^ increase; aOR:1.09, IQR:1.05-1.14, p<0.001) and presence of positive smear at baseline (aOR:1.61, IQR:1.08-2.40, p<0.001) were both independently associated with increased odds of DM but not PDM. Of note, no association was found between presence of HIV-infection and odds of presenting with PDM (p=0.956) or DM (p=0.174) in the RePORT-Brazil cohort (**Figure 4A**).

**Figure 4.**
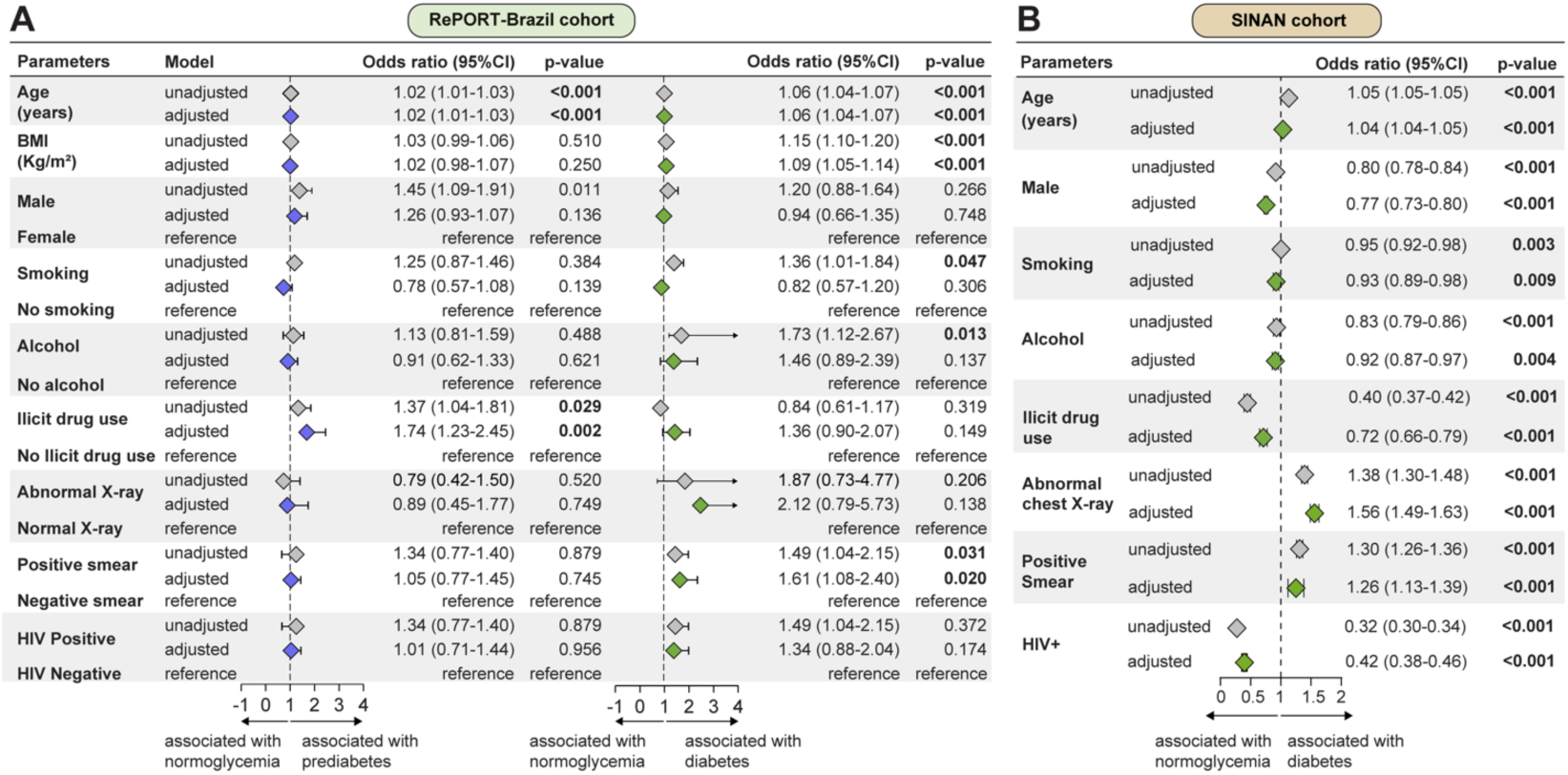
Factors associated with dysglycemia in patients with active pulmonary tuberculosis in RePORT-Brazil and SINAN cohorts. **(A)** A multinomial logistic regression analyses were used to test association between indicated characteristics of pulmonary TB patients and presence of prediabetes (left panel) or diabetes (right panel). Variables included in the adjusted model exhibited univariate p-values ≤0·2 (See **Table 2** for details). **(B)** Binomial logistic regression to test association between indicated characteristics of TB patients and presence of diabetes. Only variables with significant p-value in the adjusted model are shown. Variables included in the adjusted model exhibited univariate p-values ≤0.2 (See **Table 3** for details). Abbreviations: BMI: Body Mass Index.

To test associations between characteristics of TB and the presence of DM in the SINAN cohort, a binomial logistic regression analysis was performed. In this cohort, aging (per 1-year increase; aOR: 1.04, IQR:1.04-1.05, p<0.001), positive smear (aOR: 1.26, IQR:1.13-1.39, p<0.001) and abnormal chest X-ray (aOR: 1.56, IQR:1.49-1.63, p<0.001) at baseline were independently associated with presence of DM. In contrast, male sex (aOR: 0.77, IQR:0.73-0.80, p<0.001), current smoking (aOR: 0.93, IQR:0.89-0.98, p <0.001), alcohol consumption (aOR: 0.92, IQR:0.87-0.97, p <0.001), use of illicit drugs (aOR: 0.72, IQR:0.66-0.79, p<0.001) and to live with HIV (aOR: 0.42, IQR:0.38-0.46, p<0.001) were all associated with a decreased odds of DM (**Figure 4B**).

### 3.5 Clinical and epidemiologic profiling according to the glycemic status and HIV

In the RePORT-Brazil cohort, the TBDM-HIV, TBPDM-HIV and TB-HIV groups presented similar frequencies for male sex (≈75.9%, p = 0.002). Interestingly, the highest median age was in the TBDM group (49 years), followed by 36 years in the TBDM-HIV and TBPDM groups, with the lowest median age observed in the TB group (30 years) (p <0.001) (**Table 4**). *Pardo* race was the most reported in the TBDM-HIV group (69.6%). Drug resistance to isoniazid was more frequently observed in the groups with HIV coinfection (p = 0.036) (**Table 4**).

**Table 4.** Characteristics of TB cases by DM status in RePORT-Brazil cohort.

| Characteristics | TBDM-HIV<br>(n=56) | TBPDM-HIV<br>(n=97) | TB-HIV<br>(n=91) | TBDM<br>(n=217) | TBPDM<br>(n=341) | TB<br>(n=350) | p-value |
| --- | --- | --- | --- | --- | --- | --- | --- |
| **Sex male – no. (%)** | 43 (76.8) | 75 (77.3) | 67 (73.6) | 137 (63.1) | 232 (68.0) | 209 (59.7) | **0.002** |
| **Age – median (IQR)** | 36 (31-42) | 35 (28-43) | 34 (26-42) | 49 (38-57) | 36 (25-49) | 30 (22-42) | **<0.001** |
| **Race/Ethnicity – no. (%)** |  |  |  |  |  |  | **<0.001** |
| White | 6 (10.7) | 26 (26.8) | 11 (12.1) | 37 (17.1) | 66 (19.4) | 95 (27.1) |  |
| Black | 11 (19.6) | 12 (12.4) | 16 (17.6) | 46 (21.3) | 103 (30.2) | 106 (30.3) |  |
| Asian | 0 (0.0) | 2 (2.1) | 3 (3.3) | 4 (1.9) | 3 (0.9) | 4 (1.1) |  |
| *Pardo* | 39 (69.6) | 57 (58.8) | 61 (67.0) | 127 (58.8) | 164 (48.1) | 145 (41.4) |  |
| Indigenous | 0 (0.0) | 0 (0.0) | 0 (0.0) | 2 (0.9) | 5 (1.5) | 0 (0.0) |  |
| **BMI- (kg/m<sup>2</sup>) – median (IQR)** | 20.5 (17.5-22.5) | 20.2 (18.7-21.8) | 19.9 (17.9-22.7) | 21.9 (19.9-25.2) | 19.8 (18.1-21.8) | 19.9 (17.9-21.8) | **<0.001** |
| **Smoking – no. (%)** | 36 (64.3) | 60 (61.9) | 56 (61.5) | 120 (55.3) | 171 (50.1) | 164 (46.9) | **0.009** |
| **Alcohol consumption – no. (%)** | 53 (94.6) | 91 (93.8) | 81 (89.0) | 187 (86.2) | 271 (79.5) | 276 (78.9) | **<0.001** |
| **Illicit drug use – no. (%)** | 30 (53.6) | 47 (48.5) | 53 (58.2) | 47 (21.7) | 123 (36.1) | 85 (24.3) | **0.023** |
| **Positive smear – no. (%)** | 37 (66.1) | 58 (61.1) | 49 (55.1) | 182 (83.9) | 262 (77.3) | 271 (78.3) | **<0.001** |
| **Previous diagnosis of diabetes – no. (%)** | 9 (16.1) | 22 (22.9) | 19 (21.3) | 41 (19.2) | 52 (15.3) | 55 (15.8) | **<0.001** |
| **Abnormal chest X-ray – no. (%)** | 52 (92.9) | 81 (83.5) | 79 (86.8) | 215 (99.1) | 335 (98.2) | 344 (98.3) | **<0.001** |
| **Drug-susceptibility testing (DST) – no. (%)** |  |  |  |  |  |  |  |
| Rifampicin-Isoniazid resistance | 2 (3.9) | 5 (5.8) | 3 (4.0) | 6 (3.0) | 3 (1.0) | 7 (2.3) | 0.165 |
| Rifampicin resistance | 4 (7.8) | 5 (5.8) | 3 (4.0) | 6 (3.0) | 3 (1.0) | 10 (3.3) | 0.057 |
| Isoniazid resistance | 7 (13.7) | 12 (14.0) | 8 (10.7) | 14 (7.1) | 19 (6.3) | 16 (5.3) | **0.036** |
| Sensitive | 45 (88.2) | 77 (89.5) | 69 (92) | 177 (89.4) | 248 (82.4) | 265 (87.5) | 0.115 |
| **Symptoms of TB– no. (%)** |  |  |  |  |  |  |  |
| Hemoptysis | 6 (15.8) | 8 (10.4) | 8 (13.1) | 57 (29.1) | 78 (26.3) | 81 (27.6) | 0.175 |
| Cough | 38 (74.5) | 78 (89.7) | 61 (81.3) | 196 (99.0) | 298 (98.3) | 295 (95.8) | **0.004** |
| Fever | 45 (88.2) | 71 (81.6) | 63 (84.0) | 150 (75.8) | 240 (79.2) | 228 (74.0) | 0.846 |
| Weight Loss | 49 (96.1) | 82 (94.3) | 65 (86.7) | 185 (93.4) | 279 (92.1) | 272 (88.9) | **0.018** |
| Fatigue | 48 (94.1) | 73 (83.9) | 68 (90.7) | 165 (83.3) | 240 (79.2) | 237 (76.9) | 0.364 |
| Night sweats | 31 (60.8) | 63 (72.4) | 47 (62.7) | 142 (72.1) | 213 (70.3) | 214 (69.7) | 0.448 |
| Chest pain | 31 (60.8) | 47 (54.0) | 33 (44.0) | 134 (68.0) | 201 (66.3) | 212 (69.1) | **0.108** |

**Table note:** Data represent no. (%), except for age and BMI, which is presented as median and interquartile range (IQR). Continuous variables were compared using the Kruskal-Wallis test and categorical variables were using the Pearson's chi-square test. Bold values represent statistically significant
*Definition of alcohol consumption:* Past or current any consumption of alcohol. *Definition of smoking:* Past or current cigarette smoker. *Definition of passive smoking:* Living with someone who smokes. *Definition of illicit drug use:* Past or current illicit drug use (marijuana, cocaine, heroin or crack). *Definition of persistence of symptoms:* Patients who in the initial evaluation interview (baseline) reported indicated symptom and in the evaluation of visit 2 (month 2) still reported having such symptom. *Definition of Pardo ethnicity:* mixture of European, black and Amerindian. Abbreviations: TB: tuberculosis, BMI: Body Mass Index

TB cases with HIV comorbidity displayed lower proportions of abnormal chest radiographs (p <0.001) and of smear-positive sputum samples (p <0.001) (**Figure 5A, Table 4**). We observed that tobacco use (64.3%) and alcohol consumption (94.6%) were significantly more reported in the TBDM-HIV group when compared to the clinical groups without HIV. (**Figure 5B, Table 4**). As expected, regarding to the TB classic symptoms, the participants from the TBDM-HIV group presented a lower frequency of cough (p = 0.004) and a higher percentage of patients with weight loss (p = 0.018) **(Figure 5C, Table 4)**.

**Figure 5:**
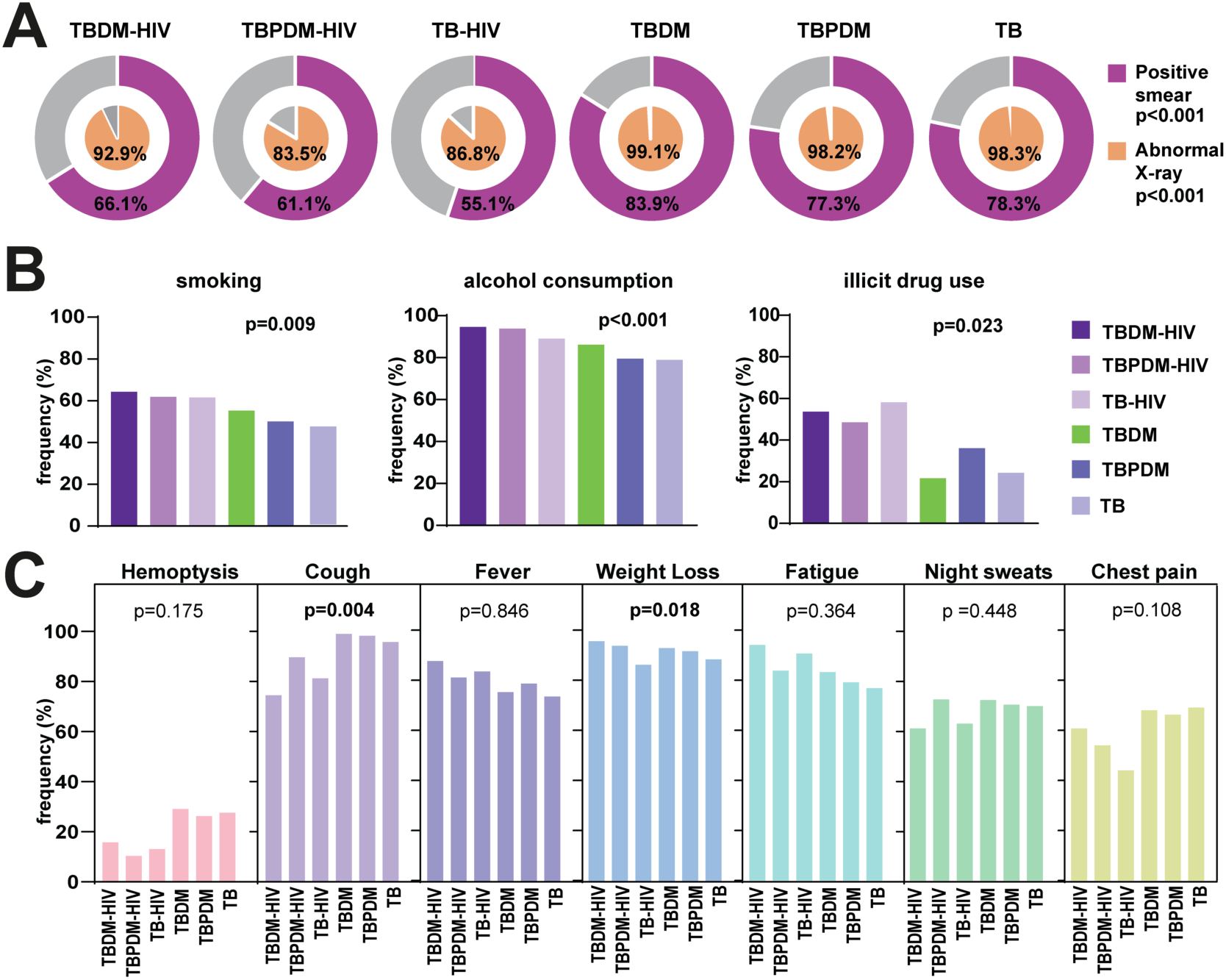
Clinical Characteristics of people with TB according glycemic and HIV status in the RePORT-Brazil cohort. **(A)** Proportion of positive smears and abnormal X-rays in each study group. **(B)** Frequency of TB cases according glycemic and HIV status regarding smoking habit, alcohol consumption and illicit drug use (smoking, alcohol, and illicit drug: in the past or at the time of evaluation before anti-TB treatment). **(C)** Frequency of TB classical symptoms in each study group. The data were compared between the groups using the Pearson’s chi-square test. Comparisons with significant p-values are displayed in bold. Abbreviations: TB: tuberculosis, DM: diabetes, PDM: prediabetes.

We found a similar clinical profile in the SINAN cohort, where the TBDM-HIV group was characterized by a higher frequency of male sex (70.1%). Furthermore, the highest median age was 55 years among TBDM cases, followed by 49 years in the TBDMHIV group (p <0.001) (**Table 5**). Such as in RePORT-Brazil, the *pardo* race was the most self-reported in all groups. Remarkably, the TBDM-HIV group presented a slight proportion of drug resistance cases, and especially to rifampicin and isoniazid (8.1%) (p <0.001).

**Table 5.** Characteristics of TB cases by DM and HIV status in SINAN cohort.

| Characteristics | TBDM-HIV<br>(n=1051) | TB-HIV<br>normoglycemia<br>(n=31073) | TB DM<br>(n=18591) | TB<br>normoglycemia<br>(n=172939) | p-value |
| --- | --- | --- | --- | --- | --- |
| **Sex male – no. (%)** | 737 (70.1) | 22339 (71.9) | 11684 (62.8) | 116122 (67.1) | **<0.001** |
| **Age – median (IQR)** | 49 (40-57) | 39 (31-46) | 55 (46-63) | 40 (28-54) | **<0.001** |
| **ART use – no. (%)** | 468 (69.1) | 13883 (72.8) | - | - | **<0.001** |
| **Race/Ethnicity – no. (%)** |  |  |  |  | **<0.001** |
| White | 341 (34.3) | 9920 (34.1) | 6220 (35.2) | 54881 (33.4) |  |
| Black/*Pardo* | 647 (65.0) | 18919 (65.0) | 11171 (63.1) | 105690 (64.4) |  |
| Indigenous | 2 (0.2) | 112 (0.4) | 150 (0.9) | 2314 (1.5) |  |
| Asian | 5 (0.5) | 169 (0.6) | 149 (0.8) | 1357 (0.8) |  |
| **Abnormal chest X-ray – no. (%)** | 772 (89.1) | 22204 (87.9) | 14824 (94.9) | 133615 (93.4) | **<0.001** |
| **Alcohol consumption – no. (%)** | 282 (28.4) | 6548 (21.5) | 2703 (14.9) | 30972 (18) | **<0.001** |
| **Illicit drug use – no. (%)** | 173 (18.5) | 5798 (19.8) | 767 (4.4) | 17305 (10.4) | **<0.001** |
| **Smoking – no. (%)** | 265 (27.9) | 6374 (21.6) | 3353 (19.1) | 33647 (20.2) | **<0.001** |
| **Positive smear – no. (%)** | 431 (61.8%) | 10937 (55.0) | 11010 (77.1) | 93512 (73.1) | **<0.001** |
| **Positive culture – no. (%)** | 195 (59.3%) | 5266 (58.3) | 3667 (65.4) | 32453 (64.3) | **<0.001** |
| **Drug-susceptibility testing (DST) –no. (%)** |  |  |  |  | **<0.001** |
| Rifampicin resistance | 4 (4.0) | 91 (3.4) | 31 (1.7) | 290 (1.8) |  |
| Isoniazid resistance | 3 (3.0) | 137 (5.1) | 95 (5.2) | 791 (5.0) |  |
| Rifampicin-Isoniazid resistance | 8 (8.1) | 119 (4.4) | 92 (5.1) | 644 (4.1) |  |
| Sensitive | 84 (84.8) | 2360 (87.2) | 1602 (88.0) | 14089 (89.1) |  |

**Table note:** Data represent no. (%), except for age, which is presented as median and interquartile range (IQR). Continuous variables were compared using the Mann-Whitney *U* test and categorical variables were using the Fisher's exact test (ART use) or Pearson's chi-square test. Bold values represent statistically significant. *Definition of Pardo ethnicity:* mixture of European, black and Amerindian Abbreviations: TB: tuberculosis, ART: antiretroviral therapy

Similar to the abovementioned results on the RePORT-Brazil, in the SINAN cohort we found a low frequency of positive smear in the TBDM-HIV (61.8%) and TB-HIV (55%) groups (p <0.001) as well as of abnormal X-rays (89.1 % and 87.9%, respectively) (p <0.001) (**Figure 6A, Table 5**). Furthermore, the positive culture results were also less frequently reported in the groups with HIV comorbidity when compared to the groups of individuals non-exposed to HIV (p <0.001). (**Figure 6B, Table 5**). Finally, TBDM-HIV cases more frequently reported the tobacco smoking (27.9%) and alcohol consumption (28.4%) (p <0.001) (**Figure 6C, Table 5**).

**Figure 6:**
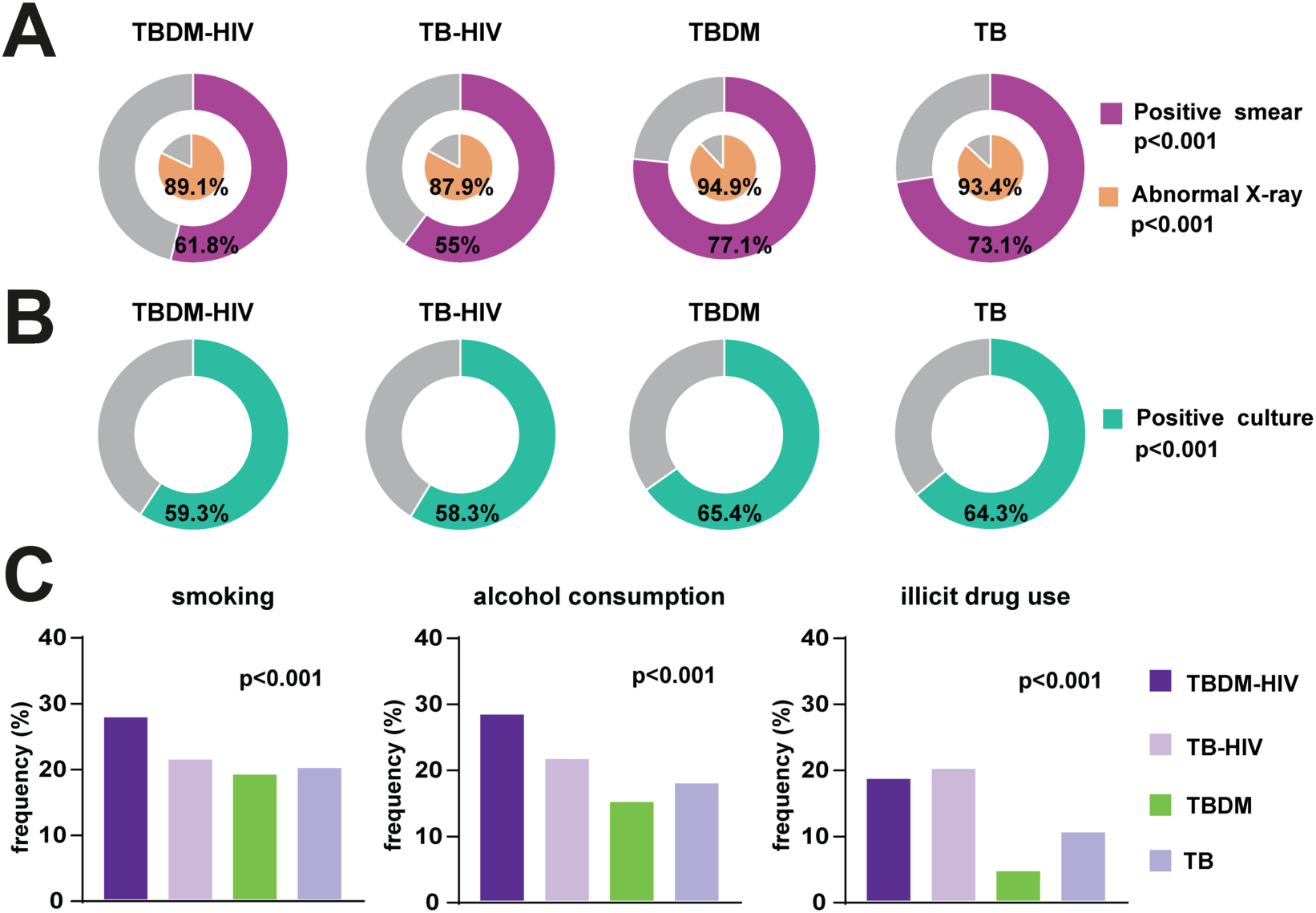
Clinical Characteristics of people with TB according glycemic and HIV status of the SINAN cohort. **(A)** Proportion of positive smears and abnormal X-rays in each study group. **(B)** Frequency of positive cultures. **(C)** TB cases according glycemic and HIV status regarding smoking habit, alcohol consumption and illicit drug use (smoking, alcohol, and illicit drug: in the past or at the time of evaluation before anti-TB treatment). The data were compared between the groups using the Pearson’s chi-square test. Comparisons with significant p-values are displayed in bold. Abbreviations: TB: tuberculosis, DM: diabetes, PDM: prediabetes.

## 4 DISCUSSION

Characterizing the association between TB, dysglycemia and HIV is important to understand the influence of metabolic and immunologic dysregulation in the presentation of the TB disease. The TB-DM association is frequent worldwide; currently, more TB patients live with DM than with HIV (Girardi et al., 2017). In the RePORT-Brazil cohort, the prevalence of dysglycemia among TB patients at baseline was 61.5% (37.8% PDM and 23.7% DM). This prevalence was higher than that recently reported in Ghana (Yorke et al., 2021), Peru (Calderon et al., 2019) and in the South of Brazil (Beraldo et al., 2021). The RePORT-Brazil cohort is large and composed by individuals from different regions of Brazil, and we have recently shown that it is representative of Brazilian patients with TB (Arriaga et al., 2021). The present work reports findings consistent with the literature, where TB-dysglycemia (mainly DM) patients have increased BMI values and higher prevalence of weight loss than normoglycemic patients (Almeida-Junior et al., 2016). TB-DM patients in RePORT-Brazil exhibited similar characteristics to those in a large cohort of 709,000 Brazilians with TB from 2007 to 2014: mostly men, mean age >40 years and self-reported black or *pardo* (Evangelista et al., 2020).

TB patients more frequently reported smoking and use of illicit drugs and alcohol, which are shared risk factors not only for TB but also for DM (Yu et al., 2014;Pelissari and Diaz-Quijano, 2018). The multinomial regression analysis demonstrated that illicit drug use was associated with increased odds of PDM, whereas alcohol use and smoking were associated with DM in the unadjusted model. Also in this analysis, aging was associated with both PDM and DM, and higher BMI was associated with presence of DM. These are factors already described as risk factors for TB in patients with DM, in addition to a lack of glycemic control (Khalil and Ramadan, 2016). The majority (55.6%) of the TB-DM patients in the RePORT-Brazil study had no previous diagnosis of dysglycemia, which can be related to a lack of glycemic control that may be contributing to a more severe symptomatology (Khalil and Ramadan, 2016), considering that coughing was a symptom associated with DM. The rate of newly diagnosed patients was high compared to other studies (Abdelbary et al., 2016), representing 66% of DM cases in RePORT Brazil cohort, demonstrating the importance of DM screening at the time of TB diagnosis.

Using data from SINAN, we observed that between 2015 and 2019, the frequency of TB-DM in Brazil was only 9.2%, lower than the global prevalence of 15% and higher than the South American prevalence of 7.7%, calculated from a meta-analysis of more than 200 studies recently conducted around the world (Noubiap et al., 2019). When comparing our original data with the results obtained through SINAN, in RePORT-Brazil, patients with TB-DM were more likely to be male, black/*pardo*, older and more frequently to have a positive sputum smear than persons without DM, reinforcing the idea that the results obtained with RePORT-Brazil are representative of the country’s population. However, in contrast to RePORT-Brazil, in SINAN, TB-DM patients had a significantly lower frequency of HIV-infection than those who did not report DM. This difference found in SINAN may be since glycemic control is performed in all the study participants diagnosed with dysglycemia, whereas it is only recommended, and not mandatory, in the national guidelines. In addition, there is a potential underreporting of cases in SINAN, and only DM cases, but not PDM, are notified. We have discussed this limitation in the SINAN database previously (Arriaga et al., 2021), where the performance of health and epidemiological indicators was substantially higher in RePORT-Brazil than in the cases notified to SINAN. In the SINAN cohort, there was a lower proportion of males in the TBDM group, probably due to the higher percentage of women diagnosed with diabetes (Abreu et al., 2020). On the other hand, the lower frequency of alcohol, smoking and illicit drug use could be attributed to the fact that this information is self-reported by patients rather than formally investigated (Santos et al., 2018).

In RePORT-Brazil, most patients with DM or PDM were HIV-seronegative. Other studies had shown this low frequency of HIV-infection in association with DM in Brazil (Almeida-Junior et al., 2016;Evangelista et al., 2020). HbA1c levels were also similar in TB patients stratified by HIV status. There is scarce evidence describing the interaction of HbA1c values and HIV in patients with TB. One study described that HbA1c could underestimate real glycemia values in PWLH (Glesby et al., 2010). Furthermore, dysglycemia risk in PLWH has been shown to be increased after initiating ART (Julius et al., 2011), which could be a potential confounding factor, but HIV-infection was not associated with occurrence of dysglycemia in our study in both cohorts, even when stratified by age. Of note, in the SINAN cohort, presence of HIV-infection was linked to increased likelihood of normoglycemia in the population with pulmonary TB. Thus, the findings presented here from both large cohorts analyzed in this study argue that HIV-infection does not appear to be a determinant of dysglycemia in patients with pulmonary TB in Brazil.

To investigate whether HIV had any influence on dysglycemia in the RePORT-Brazil cohort, we tested for correlations between HbA1c and HIV-1 viral load or CD4 T-cell counts. We found just weak and non-significant correlations, indicating that HIV progression may not influence the occurrence of significant hyperglycemia. A study in PLWH that used fasting plasma glucose to measure glycemia reported that CD4 T-cell counts, and HIV viral load could influence blood glucose levels (Duro et al., 2015). Further studies are necessary to clarify whether HIV disease progression affects glycemic control by measuring several laboratory parameters simultaneously, such as HbA1c, fasting glucose levels or oral glucose tolerance tests. Our findings clearly corroborate the idea that despite the effect of HIV-infection on the immune system, glucose metabolism does not seem to be highly affected by this infection or disease progression.

We show the groups according to the glycemic status and by HIV infection and we identified that the group of persons with TBDM-HIV present with some peculiar characteristics. Male sex, smoking and alcohol consumption were higher in the TBDM-HIV group. We did not find specific literature to be able to contrast to our results. However, a study in 132 people with HIV described that the male population has a strong association with smoking, and in turn there is a strong interaction between smoking and alcohol consumption in infected men with HIV (Bhatta et al., 2018) which is consistent with the results of our study. Immunodeficiency and a decreased inflammatory response can inhibit sputum production in individuals with HIV; such cases also tend to have fewer atypical findings on radiographs (Abaye et al., 2017), which coincides with the overall low percentage of cough and lower frequency of abnormal x-rays found in the TBDM-HIV, TBPDM-HIV and TB-HIV groups. Among the groups of individuals living with HIV, the TBDM-HIV presented a higher proportion of abnormal x-rays and self-reported cough. We hypothesize that presence of DM may boost immunopathological mechanisms that lead to tissue damage and inflammation which results in abnormal radiographs and cough. Reinforcing this idea, we have previously reported that the transcriptome of TB-DM patients exhibits increased representation of neutrophilic inflammation pathways (Prada-Medina et al., 2017), which may contribute at least in part to lung damage leading to cough and altered x-rays.

The present study has some limitations. In RePORT-Brazil, dysglycemia was investigated by means of HbA1c levels; we did not perform fasting glucose levels or oral glucose tolerance tests. Although glycated hemoglobin levels have been reliably used to estimate dysglycemia in several studies, it is possible that the final numbers of DM and PDM would have differed if additional laboratory assessments had been used. In addition, the use of anti-DM drugs was not uniformly recorded. In SINAN, diabetes condition is notified without differentiating if it was self-reported or if it had a laboratory confirmation. Therefore, the accuracy of DM diagnosis may have been affected. Regardless of its limitations, the present study adds important knowledge to the study of dysglycemia in TB patients in a large well-characterized multicenter cohort from Brazil, enabling the identification of factors associated with PDM and DM in this population. We also demonstrate that the majority of patients with TB-DM had no previous diagnosis of dysglycemia, which may be associated with an underreporting of DM in the SINAN database, and that HIV-infection was not significantly associated with dysglycemia in TB patients. It is important to systematically screen for DM in TB patients and initiate appropriate therapy for both diseases to reduce the dual burden of these major diseases.

## Data Availability

All data produced in the present study are available upon reasonable request to the authors.

## 7 Author Contributions

Conceptualization, T.R.S., M.C.F., M.C.S., V.C.R., and B.B.A.; Data curation, M.B.A., M.A-P., A.T.L.Q., M.M.S.R., and B.B.A.; Investigation, M.B.A., M.A-P., B.B-D., C.S., J.P.M.P.,E.V.N., B.M.F.N., M.S.R.,A.B.S.,A.B., J.G.O., A.S.R.M., R.S.G., M.C.F., B.D., J.R.L.S., A.L.K., S.C., V.C.R., T.R.S., M.C.S., and B.B.A.; Formal analysis, M.B.A., M.A-P., B.B-D. and B.B.A.; Funding acquisition, B.D., J.R.L.S., A.L.K., S.C., V.C.R., T.R.S., M.C.S., M.C.F., and B.B.A.; Methodology, M.B.A., M.A-P., B.B-D., and B.B.A.; Project administration, M.C.F., T.R.S., and B.B.A.; Resources, M.B.A., M.A-P., B.B-D., T.R.S., and B.B.A.; Software, M.B.A., M.A-P., A.T.L.Q., M.M.S.R., M.C.F., T.R.S., and B.B.A.; Supervision, T.R.S., and B.B.A.; Writing—original draft, M.B.A., M.A-P., B.B-D., J.P.M.P., C.S., E.V.N. and B.B.A.; Writing—review and editing, all authors. All authors have read and agreed to the submitted version of the manuscript.

## 9 Acknowledgments

The authors thank the study participants. We also thank the teams of clinical and laboratory platforms of RePORT-Brazil. A special thanks to Elze Leite (FIOCRUZ, Salvador, Brazil), Eduardo Gama (FIOCRUZ, Rio de Janeiro, Brazil), Elcimar Junior (FMT-HVD, Manaus, Brazil), and Hilary Vansell (VUMC, Nashville, USA) for administrative and logistical support.

## 10 Contribution to the Field Statement

Persons with dysglycemia (i.e., diabetes or prediabetes) have increased odds of becoming infected with tuberculosis and of progressing from latent to active disease. Moreover, people with tuberculosis-diabetes comorbidity exhibit increased risk of unfavorable outcomes and mortality during anti-tuberculosis therapy. This context is complicated by the presence of another known epidemic that is HIV. The comorbidity of tuberculosis-HIV accelerates immune deterioration and represents the highest burden of infectious diseases in low-income countries. There are scarce investigations addressing the interaction between diabetes, tuberculosis, and HIV. In the present study, we examined two large cohorts of persons with pulmonary tuberculosis in Brazil to demonstrate that patients with tuberculosis-dysglycemia present a distinct clinical profile, being mainly older and presenting a greater severity of symptoms than individuals with normoglycemic tuberculosis. Moreover, our results indicate that the presence of HIV does not substantially affect the clinical presentation of people with tuberculosis-dysglycemia, although it is associated with a more frequent use of recreational drugs and negative sputum samples during tuberculosis screening. The findings provide robust evidence to support bidirectional screening of tuberculosis and diabetes. Future studies are warranted to investigate whether HIV impacts clinical presentation and outcomes of persons with tuberculosis-diabetes comorbidity in other highly endemic regions.

